# Alexander Disease: Screening and *in silico* functional characterization of variants on candidate gene

**DOI:** 10.1101/2024.02.19.24302650

**Authors:** Surajit Malakar, Tathagata Das, Vikrant Yadav, Anurag Sahu, Parimal Das

## Abstract

**Background:** Alexander disease is a rare neurological ailment caused by the degeneration of the white matter of the brain accompanied with the formation of Rosenthal fibers, a unique cytoplasmic inclusion within astrocytes (non-nerve tissue) in the brain. Till to date only heterozygous *de novo* mutation in Glial Fibrillary Acidic Protein (*GFAP*) gene is found to be associated for the disease phenotype.

**Methods and Result:** the aim of our study was to determine the genetic basis of an Indian-origin juvenile AxD patient with pathological symptoms of macrocephaly and psychomotor delay followed by regression, spastic parapresis and feeding difficulty. The patient was screened for mutation in candidate gene for AxD by Whole Exome Sequencing and the detected variants were further reconfirmed by Sanger sequencing. The clonicopathological feature were investigated and two heterozygous missence variants (c.983T>C and c. 626G>A) were indentify in GFAP gene of the patient. Familial screenings of both of these variants were predicted to be pathogenic or damaging by various *in silico* methods and prediction tools.

**Conclusion:** This altered GFA protein get accumulated in cytoplasm of the astroglial cells, leading to formation of Rosenthal fibers, which impairs cell function. Future study, however, would be helpful to understand the functional mechanism of these variant in formation of Rosenthal fibers leading to AxD pathophysiology.

## Introduction

Alexander disease (AxD) (MIM#203450) is a rare neurological condition affects the central nervous system,^1^ with an incidence rate of 1 in 2.7 million individuals globally^2^. The pathological hallmarks of the disease is characterized by the presence of Rosenthal fibers in the astrocytes as a consequence of abundant accumulation of cytoplasmic inclusion composed of Glial Fibrillar Acidic Protein (GFAP), heat shock protein 27 and αβ-crystalline^13^. This cytoplasmic inclusion affects the myelination of the nerves which predominantly affects infants and children and can also affects adults^4^.

Still to this day Glial Fibrillary Acidic Protein (*GFAP*) is the potential candidate gene responsible for AxD pathology, inheritated in autosomal dominant pattern^5,6^. GFAP is a type III intermediate filament protein transcribe from a 10 kb region on chromosome 17q21^7,8^. The gene is conserved, existing as a single copy found in both the human and rodent genomes^9,10^. It produces the primary protein product α-GFAP, which shares 91% amino acid identity (95% similarity) between humans and mice^11^. GFAP exhibits alternative splicing; however, the primary splice variant, known as α-GFAP, is mostly expressed in central nervous system astrocytes^1^. Depending on age of onset AxD was classified as infantile (onset before 2years of age), juvenile (onset 2-4 years of age) and adult (late juvenile to adult). However, on finding overlaps between the age dependent subtypes especially between juvenile (onset 2-14 years of age) and adult (late juvenile to adult) a revised classification has divided AxD into two subtypes: Type1 and Type2^2^. The infantile type I AXD are more frequent and accounts 51 % of the total AXD cases, these appears before 2 years of age and the children can live anywhere from a few days after birth to around the age of 8 to 10 years^12^. The physiological condition of juvenile AXD includes macrocephaly and psychomotor delay followed by regression, spastic parapresis and feeding difficulty. MRI is the only diagnostic test available for this condition, which reveals significant alterations in cerebral white matter, with a frontal preponderance and a comparative sparing of occipital and temporal white matter. In certain cases, it also shows abnormalities in the basal ganglia and thalamus^13^.

In the present study we have analyzed a novel (c.983T>C) and a known (c. 626G>A) missense variation at sixth and fourth exon on *GFAP* in the proband between 0-2 years of age in heterozygous condition respectively. The known missense variation (c.626G>A) was also found in phenotypically normal mother in heterozygous condition. The *In-silico* analysis of the altered protein (p.L328P and p.R209Q) predicts the structural and functional instability leading to disease pathology. This altered GFAP protein get accumulated in cytoplasm of the astroglial cells, leading to formation of Rosenthal fibers, which impairs cell function and responsible for disease phenotype.

## Material and methods

### Clinical assessment and genetic analysis

The present study involves an infantile AxD male patient of age within 0-2 years. Complete neuronal evaluations including MRI, CT scan examination was conducted. 10 ml of intravenous blood sample was collected from the proband and his parents in a heparinized vial after receiving informed consent from the parents as approved by Institutional Ethics Committee. Genomic DNA was isolated using standard salting-out protocol. Initially the proband was chosen for genetic investigation using WES.

### Whole Exome Sequencing and validation of identified variant by Sanger sequencing

WES was performed with 1.0μg of genomic DNA sample from Life Cell Biology Lab Ltd. Reconfirmation of the mutation was done by Sanger sequencing. For performing Sanger Sequencing, forward and reverse intronic PCR primers were designed using Primer3plus software. PCR was carried out using 25-50ng of DNA in ABI 96 well thermo cycler (Applied Biosystems, USA) and were purified by Exonuclease 1 and recombinant Shrimp Alkaline Phosphate (Thermo fisher scientific). Sequencing of the purified PCR products was done after labelled with ABI Big Dye Terminator V3.1 cycle sequencing kit followed by automated sequencing in ABI 3500 Genetic Analyzer according to manufacture protocol.

### Bioinformatics analysis

#### Pathogenicity prediction

Different algorithm-based online tools were used to detect the pathgenicity of the variant. SIFT, Polyphen 0.2, Mutation taster online tools were used. Depending on require inputs from the user and have a need for the mutation specific information, the outputs are based on the threshold values and the cut-off values^14,15,16,17^.

### Protein stability prediction

The impact of mutation on GFAP variants’ on protein stability was calculated by Gibbs free energy algorithms by a plethora of tools including Cupsat^18^ (Cologne University protein stability analysis tool), mCSM^19^ (mutation cutoff scanning matrix), SDM^20^, Dynamut^21^ and DUET^19^. All these tools provided ddG values along with a score that indicate whether the GFAP variations under evaluation were stabilizing or destabilizing.

### Protein structure prediction

3D protein structure has been predicted by reassembling structural fragments from threading templates using replica exchange Monte Carlo simulations by iterative threading assembly refinement algorithm (I--TASSER) ^22^.

### Physiochemical analysis

The physiochemical properties of the wild type GFAP and the mutated varients were studied by ProtScale and protparam server^23^. It detects, computes and shows the specific profile of an amino acid on a given protein, and delivered the result on hydropathicity, hydrophobicity, polarity, alpha helix and beta-sheet depending on each variant, and their respective changes.

### Post Translational Modification (PTM)

The functional diversity of the wild and the mutant type of GFAP protein molecule in presence of various functional group were investigated by various PTM software including Netphos 3.1^24^ (Phosphorylation), NetGlyc 1.0, NetOGlyc 4.0^25^ (glycosylation), GPS-SUMO^26^ (sumoylation), GPS-SNO^27^ (S-nitrosylation), GPS-PAIL (N-acetylation)^28^, PrePs^29^ (Prenylation), GPS-Palm ^30^(Palmitoylation), GPS-Uber (Ubiquitination)^31^, KprFunc^32^ (Propinylation) and GPS-Msp^33^ (Methylation). It is necessary for understanding the PTM, which eventually puts a transparent view in disease pathogenicity depending on various functionalities of the protein.

### Molecular simulation

The conformational transition of wild type and mutant variant of GFAP was studied by GROMACS software. For this study the GROMOS9643a1 forcefield was used, considering the protein model soluble in water. The framework utilized the steepest descent approach to minimize the parameters of energy, equilibrated at 310 Kelvin temperature, 1 bar pressure using the canonical ensemble (NVT) (constant number of particles, temperature, and volume) and the (NPT) outfit (constant number of particles, temperature and pressure)^34^.

Additionally other parameters including MD simulation time raised to 50ns, MD integrator Leap-frog and 5000 frames were maintained per simulation. Root-mean square deviation (RMSD), Root-mean square fluctuation (RMSF), Radius of gyration (RoG), H-bonds between protein and water, Surface accessible solvent area (SASA), were all calculated and plotted^35^.

## Result

### Whole Exome Sequencing and validation of identified variant by Sanger sequencing

WES data revealed >66,000 variant in the proband. One novel non-synonymous mutation at the position c.983 T>C (p. L328P, NM_002055.5) and a non-synonymous known mutation at the position c.626G>A (p. R209Q, NM_002055.5, rs771699440) was found on *GFAP*, the candidate gene responsible for the Alexander Disease. Both these variants were in heterozygous condition and were reconfirmed by Sanger sequencing in the proband and in the parents. The mother was found positive in carrying the known mutation c. 626 G>A and in heterozygous condition.

### Pathogenicity prediction

Various *in silico* prediction tools were used for pathogenicity prediction of both the variants on GFAP. Herein, the novel variant c.983 T>C falls under the pathogenic criteria and the known variant c. 626 G>A as likely pathogenic. The detail is mentioned in the table 2:

**Table 1.** *GFAP* variants found in Indian AxD patient.

| Nucleotide change | Location | Amino Acid change | Type of change | Allele Frequency (%) | Status |
| --- | --- | --- | --- | --- | --- |
| c. 983T>C | Exon 6 | p.L328P | Non-synonymous | NA | Novel |
| c.626G>A | Exon 4 | p.R209Q | Non-Synonymous | 0.002 | rs771699440 |

**Table 2:**
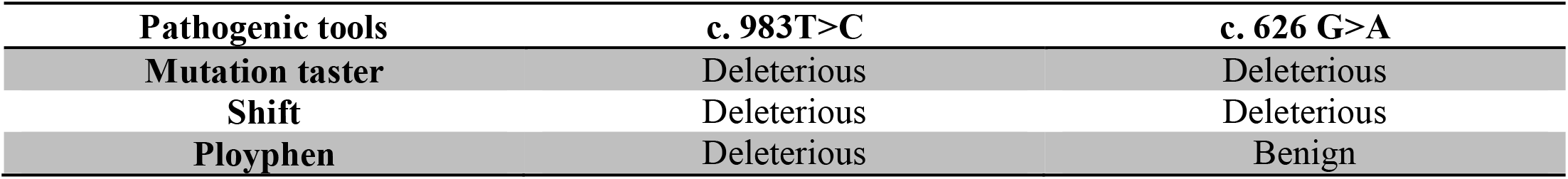
Pathogenicity prediction of GFAP c.983T>C and c. 626 G>A variants by different *In silico* tools.

### Protein stability prediction

5 popular bioinformatics prediction tools were used to predict the GFAP variants using their protein sequences, mutational positions and amino acid residues. The detail is mentioned in the table 3:

**Table 3:** Protein stability of *GFAP* variants.

| Amino acid change | Cupsat (Kcal/mol) | mCSM (Kcal/mol) | SDM (Kcal/mol) | Dynamut (Kcal/mol) | DUET (Kcal/mol) |
| --- | --- | --- | --- | --- | --- |
| p.L328P | -1.31<br>Destabilizing | -0.687<br>Destabilizing | -1.56<br>Destabilizing | -0.381<br>Destabilizing | -0.704<br>Destabilizing |
| p.R209Q | -0.39<br>Destabilizing | -0.265<br>Destabilizing | -0.07<br>Destabilizing | 0.214<br>Stabilizing | 0.014<br>Stabilizing |

### Physiochemical analysis

The physiochemical properties of the protein are very well influenced by its sequence and the characteristic component of the amino acid residues. The secondary and tertiary structures of proteins, as well as functional domains and motifs, are all determined by the characteristics of amino acid residues. As a result the protscale server was used to estimate the influence of *GFAP* mutant variations on the physio-chemical characteristic in respect to the wild type protein. Figure depicts the hydrophobicity profile and other characteristics of the wild type and GFAP mutant proteins.

### Evaluation of modeled structures

I-trasser and Saves server was used to Predict and evaluate all the mutant and wild type protein structures. The structure of highest confidential score and more than 80% of residues fall into highly favored areas in Ramachandran plot was selected.

### Molecular Dynamics Simulation

### Post translational Modification

The tables analyze the post translational modifications of the GFAP mutant proteins compared against the wild type protein. The investigation revealed Glycosylation getting affected due to the variants. As a consequence, five important protein functions involving protein interaction, activity, stability, trafficking and thermodynamics were compromised. Cellular signaling and protein stability were affected due to L328P and the double mutant changes, when analyzing tyrosine nitration. Similarly, phosphorylation was impacted due to L328P and the double mutant, leading to improper protein-protein interaction and protein trafficking.

## Discussion

Alexander Disease was first describe by W.S Alexander, is rare but often fatal neurological disorder that affects the white matter of the central nervous system, or leukodystrophy^2^. Based on the age of onset Alexander disease was formerly divided into infantile, juvenile and adult forms. However revised classification, distinguishes Alexander disease into two types: type1 and type2. Type 1 is the early onset and most severe form of Alexander disease predominantly characterized by: seizures, macrocephaly, motor and developmental delay, failing to thrive Paroxysmal degeneration, encephalopathy and typical brain MRI. Type 2 is present across the life span and has predominant features such as: eye movement abnormality, palatal clonus, bulbar involvement, autonomic dysfunction cognitive and other neurological deficiencies^12,13^.

Neuropathologically AxD is characterized by the presence of Rosenthal fibers. These Rosenthal fibers are unique cytoplasmic inclusion found within astrocytes and contain GFAP, a major astrocytic intermediate filament protein, like other cytoplasmic intermediate filament proteins. GFAP found in individual filaments and bundles within cytoplasm, passing from the perinuclear region to the periphery of the cell. In the present study, whole Exome Sequencing identified a missence, non-synonymous novel variant in exon 6 of *GFAP*, along with another non-synonymous, missence variant in exon 4 of same gene in the proband (Table 1). Sanger sequence analysis of the parents confirms the presence of the missence variant on exon 4 in the mother (Fig 2). MRI and other clinical diagnosis confirm the AxD pathophysiology in the patient whereas the mother was clinically normal.

**Fig 1:**
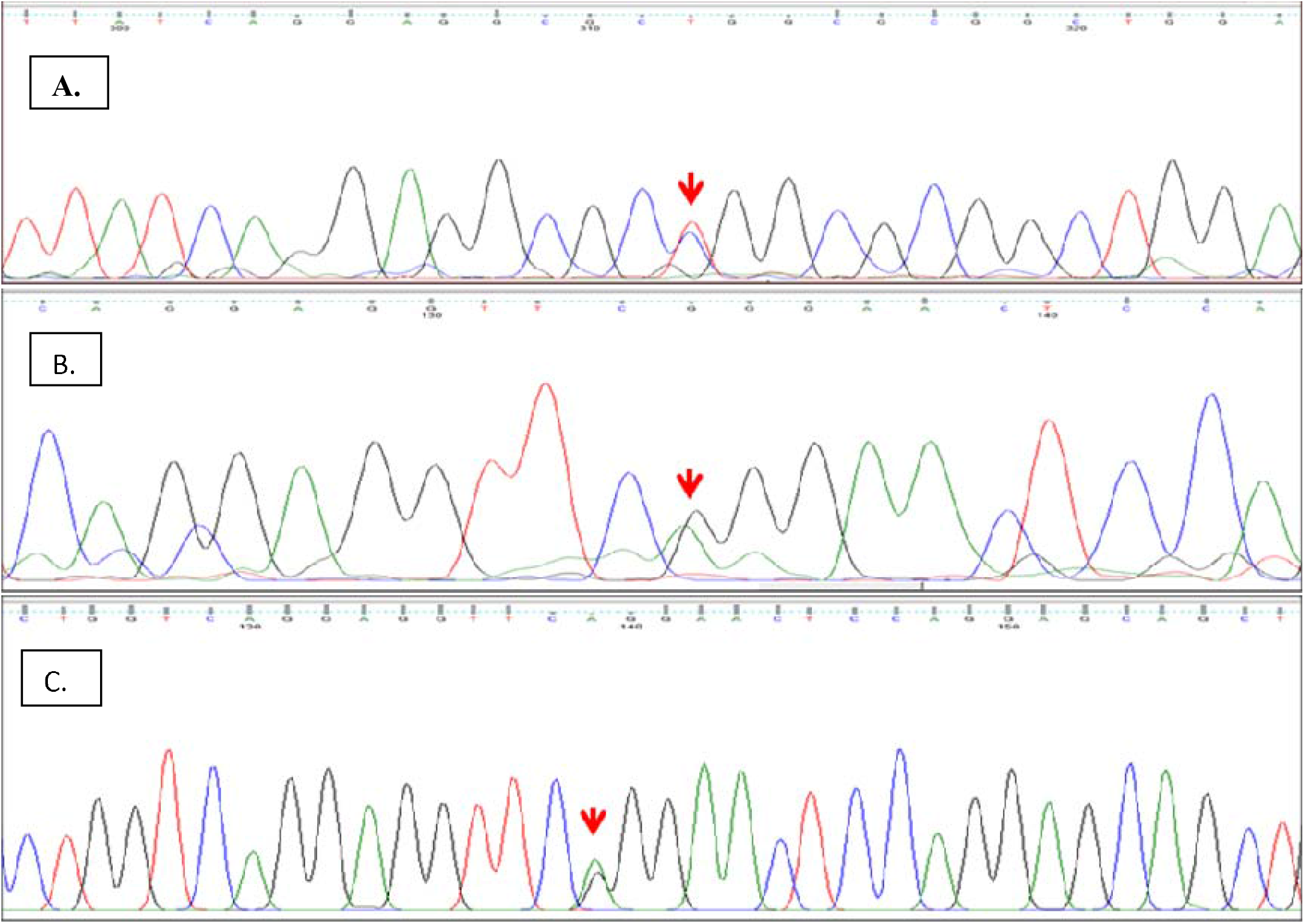
Representative DNA electropherograms of *GFAP* (A) AxD patient chromosome harboring the novel p.L328P variant due to c.983T>C heterozygous transition (B) AxD patient chromosome harboring the p.R209Q variant due to c.626G>A heterozygous transition (C) mother chromosome harboring the p.R209Q variant due to c.626G>A heterozygous transition.

**Fig 2:**
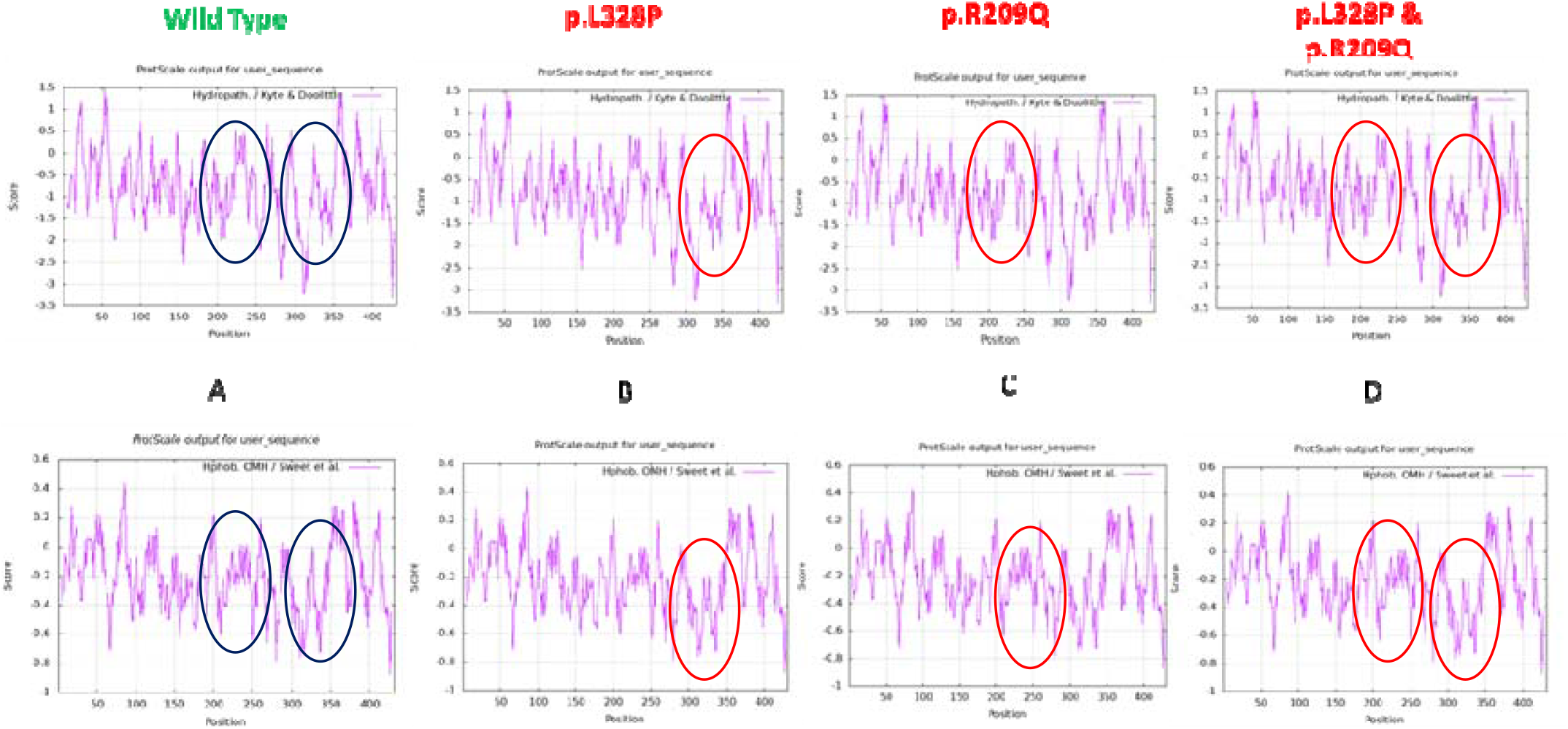
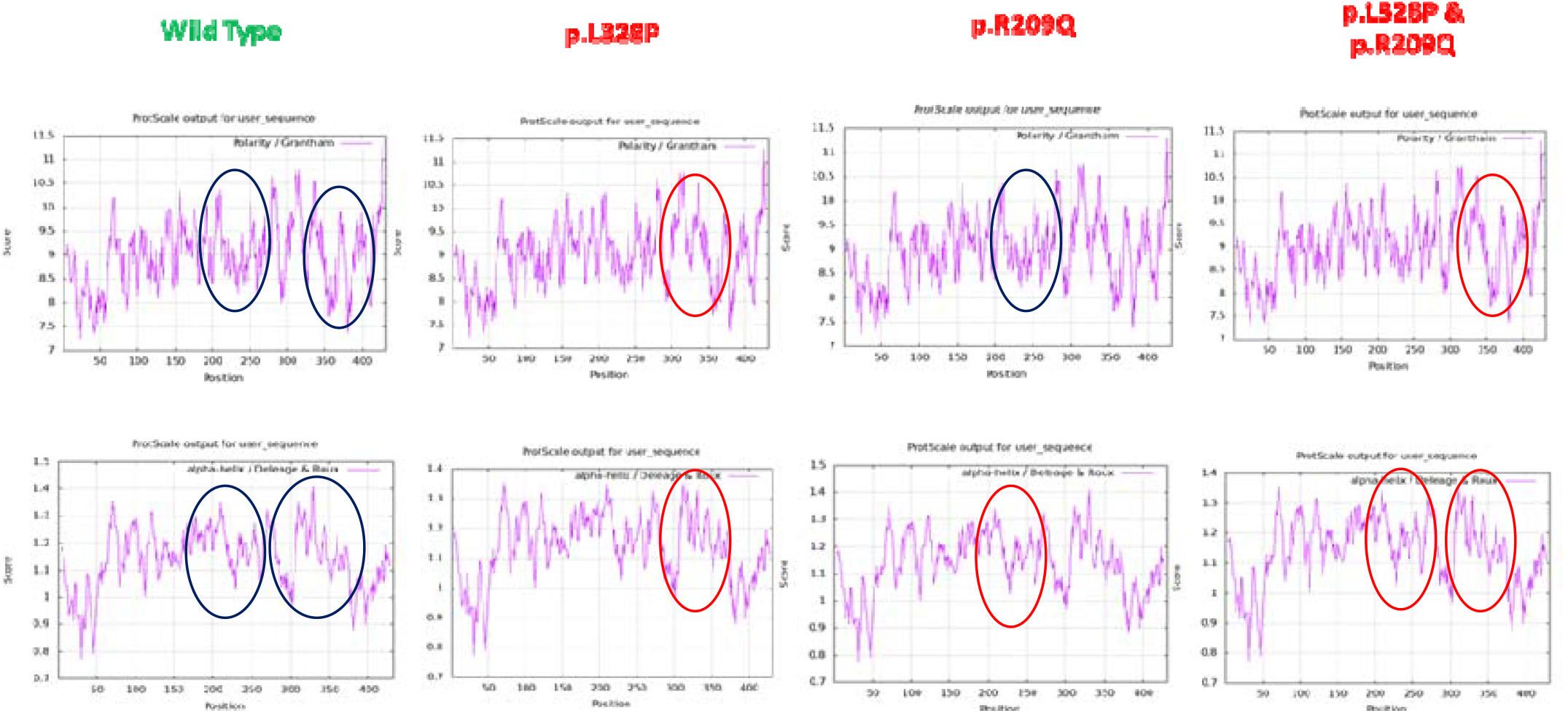
Physiochemical properties of *GFAP* wild *vs* Mutant types (P1: p.L328P, P2: p.R209Q and DM: p.L328P & p.R209Q) (A-D) Denotes Hydropathicity, (E-H) denotes Hydrophobicity, (I-L) denotes Polarity, (M-P) denotes Alpha helix. Blue circle represent wild type/ no change and red circle represent change in the plot

Previous studies have found that mutation in *GFAP* sequence interfere with some step in the polymerization process of GFAP, perhaps resulting in a misfolded protein that engages in aberrant interactions with other cellular components, compromising normal cell function^36^. The presence of ubiquitin in the Rosenthal fibers and the upregulation of the chaperones-heat shock protein 27 and αβ-crystallin, indicate that the cell perceives the aggregated GFAP as non-native and is making a valiant but ineffective attempt to break it down^1^. With sustained presence, the filament generated develops into the masses, observed enmeshing the Rosenthal fibers compromising the normal function of the astrocytes. In addition the normal mitochondrial function of the astrocytes might be affected exhibiting oxidative stress and abnormalities in ultra structure of mitochondria near Rosenthal fiber have been noted^37,38^.

Majority of the *in silico* prediction tools like mutation taster, ployphen and shift predicted these variant to be disease causing and highly deleterious (Table 2). The missence variant p. L328P on exon 6, has not been found in population till to date, and is novel to the best of our knowledge. Another missence variant p.R209Q on exon 4, with a MAF of 0.002 (Global) and 0.004 (Asian) respectively has been previously identified in European, American and in Asian population but first to be identified in Indian population by us in both the patient and his mother genome. Protein stability tools predict p. L328P variant to be more destabilizing than p. R209Q vaiant (Table 3). A significant change in the physiochemical properties (hydrophobicity, hydropathicity, alpha helix, polarity and molecular weight) was observed between the p. L328P mutant protein (P1), p.R209Q mutant protein (P2) and the double mutant protein (p.L328P + p.R209Q) (DM) repectively (Fig 3, Table 4). Significant change in tertiary protein structure of DM was found in comparison with P1, P2 and wild, which is further sustain by RMSD, RMSF, Radius of Gyration plots, Solvent Accessible Surface plots and Hydrogen bonding plots obtain from molecular dynamics simulation results (Fig 6). Principal component analysis helps in the easy visualization of complex data. It investigates key principle characteristics and components that influence the major outcomes. PCA reduces the dimensionality of data and makes it easy to interpret. PCA plots above shows the correlation of the wild type with the variants. The cosine angle in between the vectors represents the coefficient of correlation between the variants (Fig 7). Further detail about PCA can be referred from https://doi.org/10.1098/rsta.2015.0202; https://doi.org/10.1016/B978-0-12-815739-8.00012-2. PCA is used to visualize the data in an exquisite way and for a better understanding of MD simulations of the wild type and the mutant variants.

**Table 4:**
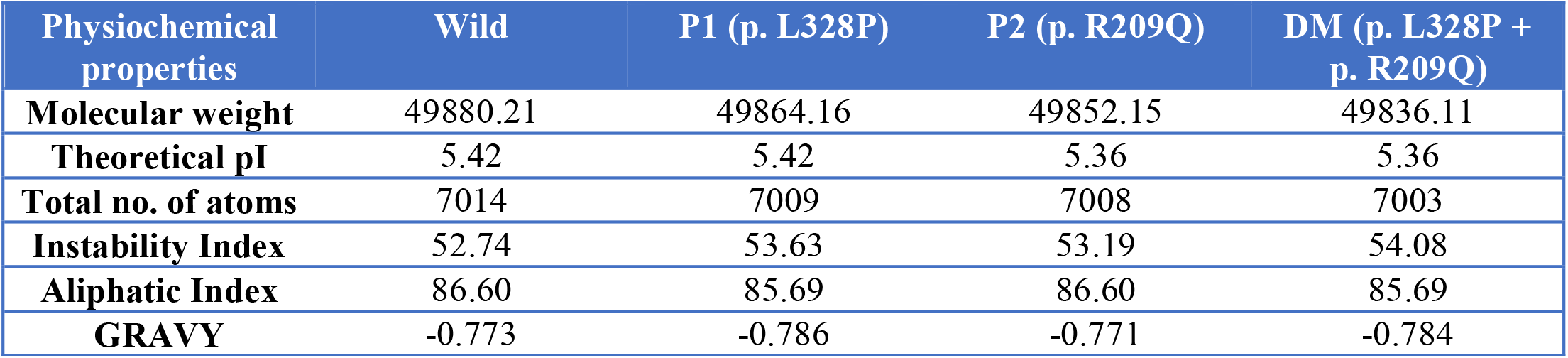
Difference in physiochemical properties of *GFAP* wild type and mutant variants.

| Physiochemical properties | Wild | P1 (p. L328P) | P2 (p. R209Q) | DM (p. L328P + p. R209Q) |
| --- | --- | --- | --- | --- |
| **Molecular weight** | 49880.21 | 49864.16 | 49852.15 | 49836.11 |
| **Theoretical pI** | 5.42 | 5.42 | 5.36 | 5.36 |
| **Total no. of atoms** | 7014 | 7009 | 7008 | 7003 |
| **Instability Index** | 52.74 | 53.63 | 53.19 | 54.08 |
| **Aliphatic Index** | 86.60 | 85.69 | 86.60 | 85.69 |
| **GRAVY** | -0.773 | -0.786 | -0.771 | -0.784 |

**Table 5:**
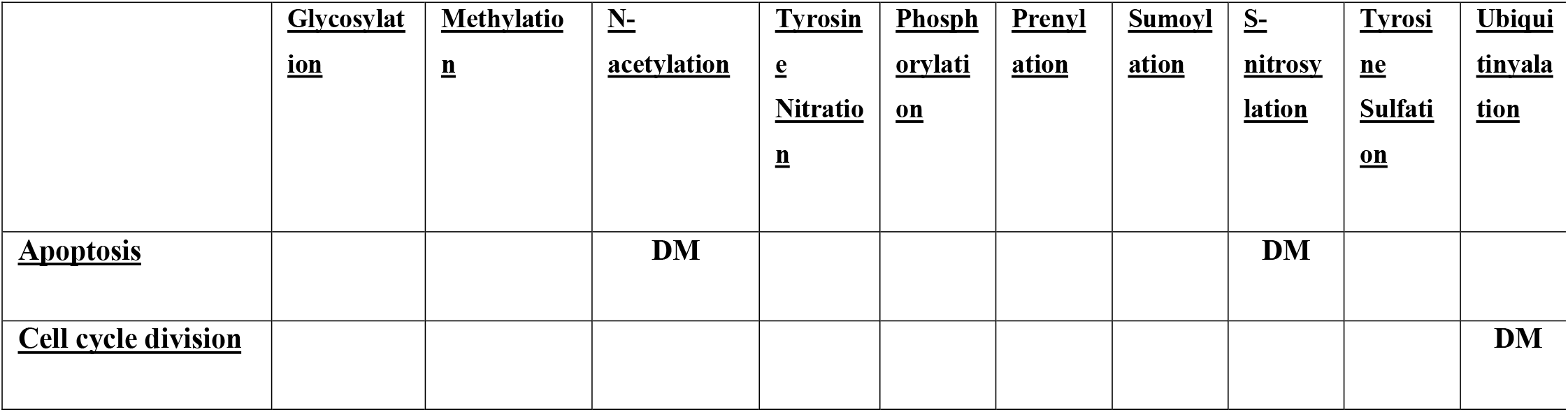
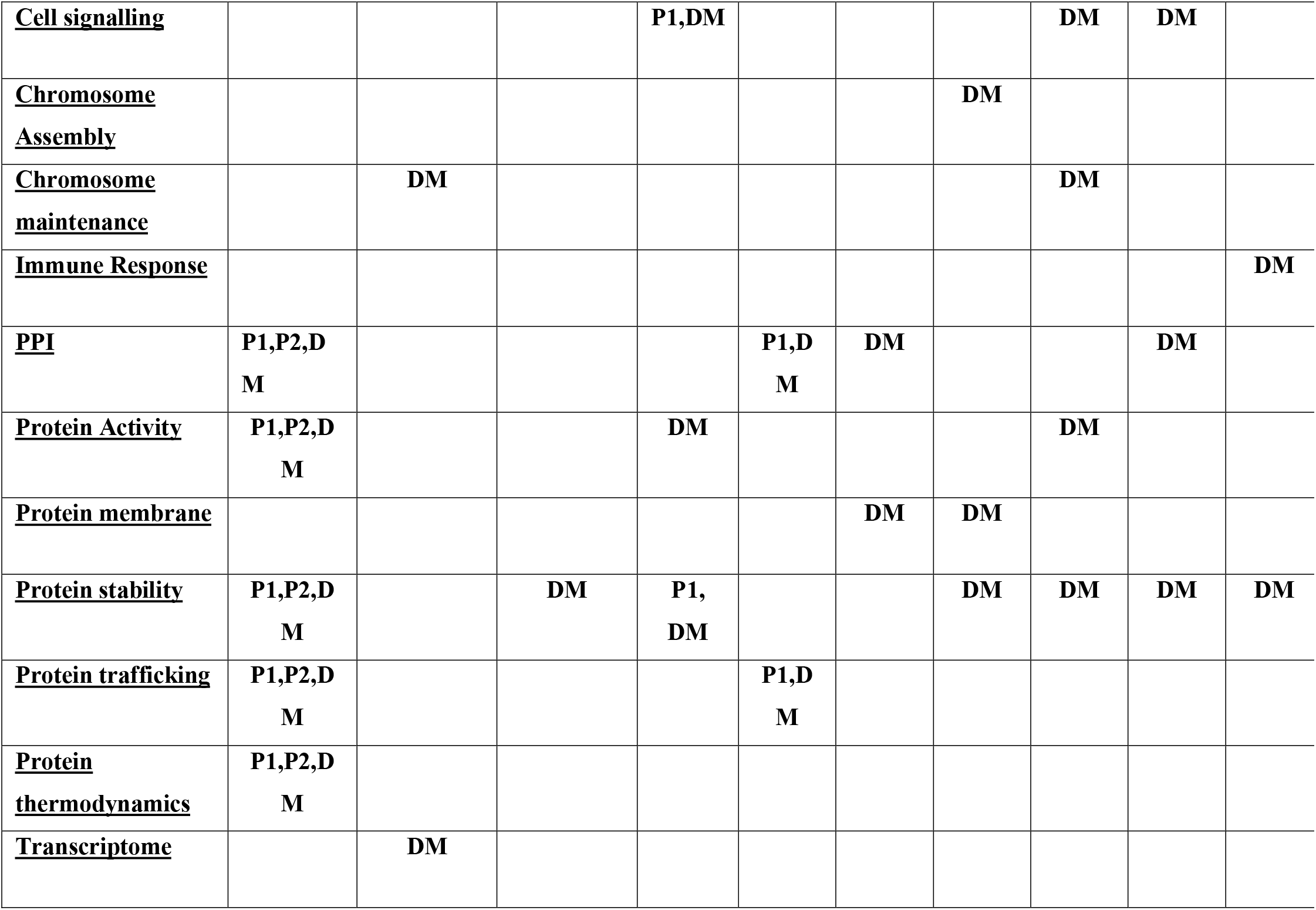
Table: Impacts on the Post-translational modification due to the mutations in the protein. The variants responsible for changes in the respective functional aspect are mentioned. (P1: p. L328P, P2: p. R209Q, DM (Double Mutant): p.L328P + p.R209Q)

|  |  |  |  |  |  |  |  |  |  |  |
| --- | --- | --- | --- | --- | --- | --- | --- | --- | --- | --- |
| **<u>Cell signalling</u>** |  |  |  | **P1,DM** |  |  |  | **DM** | **DM** |  |
| **<u>Chromosome Assembly</u>** |  |  |  |  |  |  | **DM** |  |  |  |
| **<u>Chromosome maintenance</u>** |  | **DM** |  |  |  |  |  | **DM** |  |  |
| **<u>Immune Response</u>** |  |  |  |  |  |  |  |  |  | **DM** |
| **<u>PPI</u>** | **P1,P2,DM** |  |  |  | **P1,DM** | **DM** |  |  | **DM** |  |
| **<u>Protein Activity</u>** | **P1,P2,DM** |  |  | **DM** |  |  |  | **DM** |  |  |
| **<u>Protein membrane</u>** |  |  |  |  |  | **DM** | **DM** |  |  |  |
| **<u>Protein stability</u>** | **P1,P2,DM** |  | **DM** | **P1,DM** |  |  | **DM** | **DM** | **DM** | **DM** |
| **<u>Protein trafficking</u>** | **P1,P2,DM** |  |  |  | **P1,DM** |  |  |  |  |  |
| **<u>Protein thermodynamics</u>** | **P1,P2,DM** |  |  |  |  |  |  |  |  |  |
| **<u>Transcriptome</u>** |  | **DM** |  |  |  |  |  |  |  |  |

**Fig 3:**
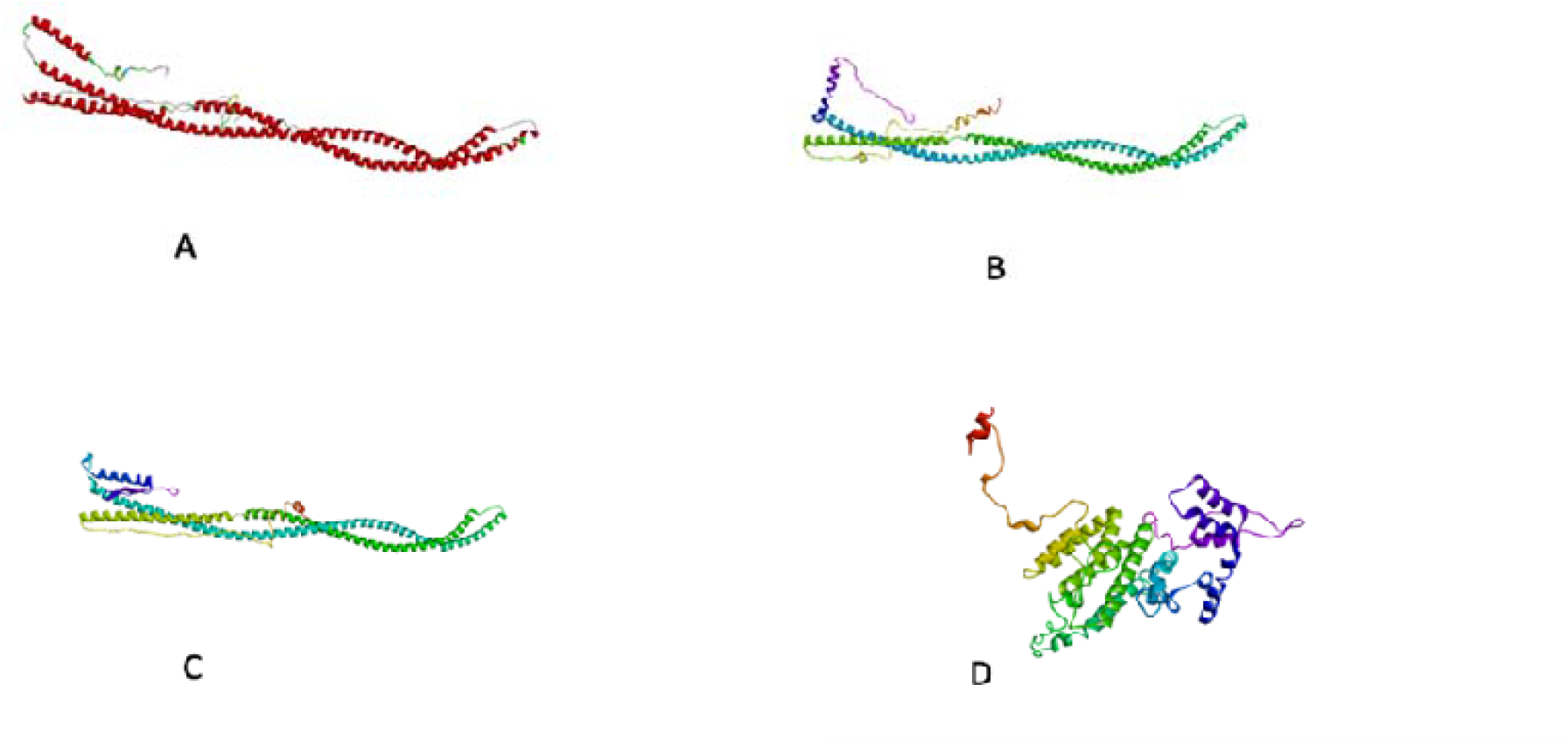
Prediction of GFAP protein model (A) Wild type, (B) P1: p.L328P (C) P2: p.R209Q (D) DM: p.L328P + p.R209Q

**Fig 4:**
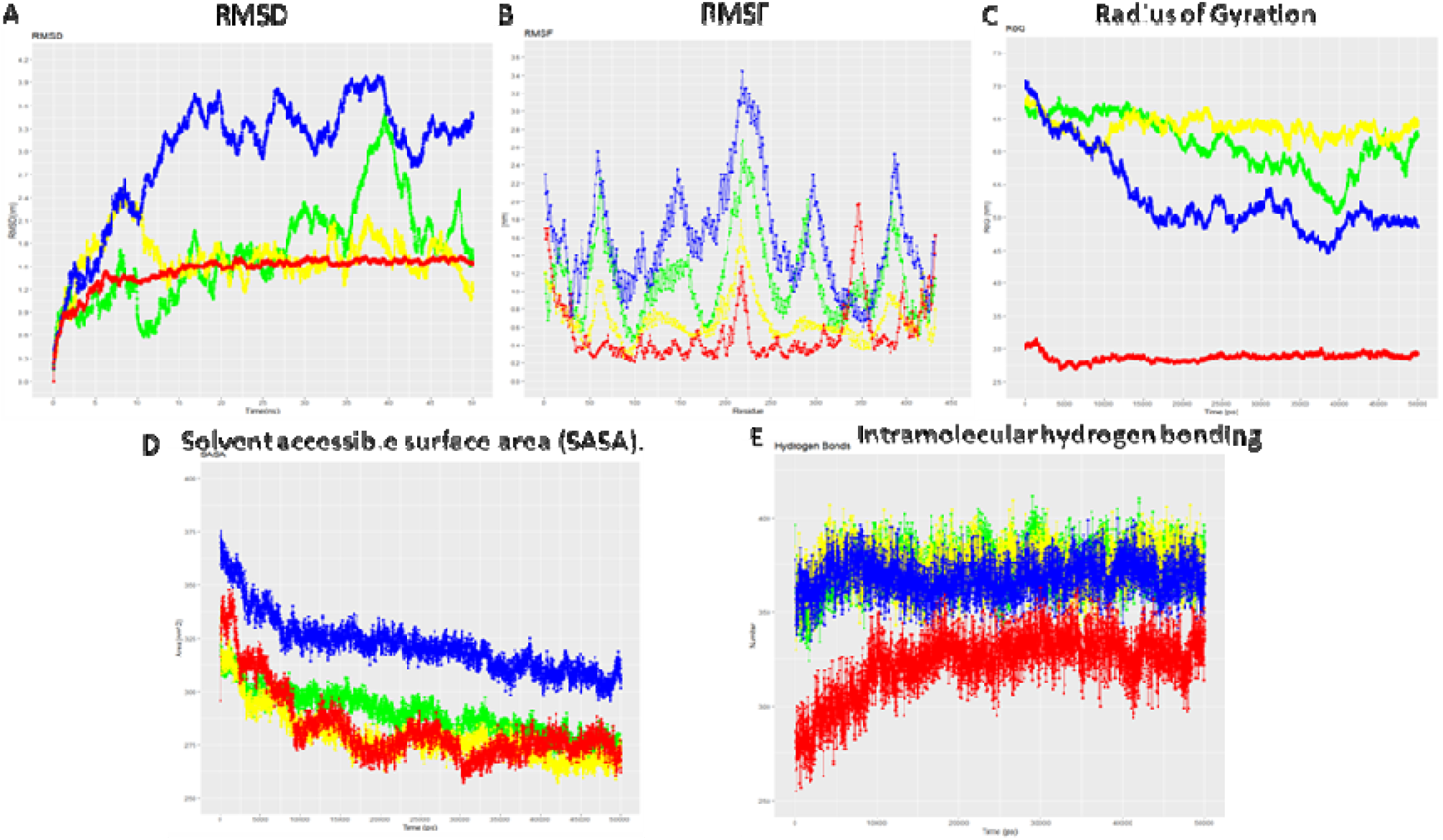
Molecular simulation results of the wild type and mutant variants (P1: p.L328P, P2: p.R209Q and DM: p.L328P & p.R209Q) at 50 ns time period. (A) RMSD plots. (B) RMSF plot. (C) Radius of gyration (Rg). (D) Solvent Accessible Surface Area (SASA). (E) Intramolecular hydrogen bonding. Wild type- Green, p.R209Q- Yellow, p.L328P- Blue and both p.L328P & p.R209Q -Red. RMSD: Root Mean Square Deviation; RMSF: Root Mean Square Fluctuation

**Fig 5:**
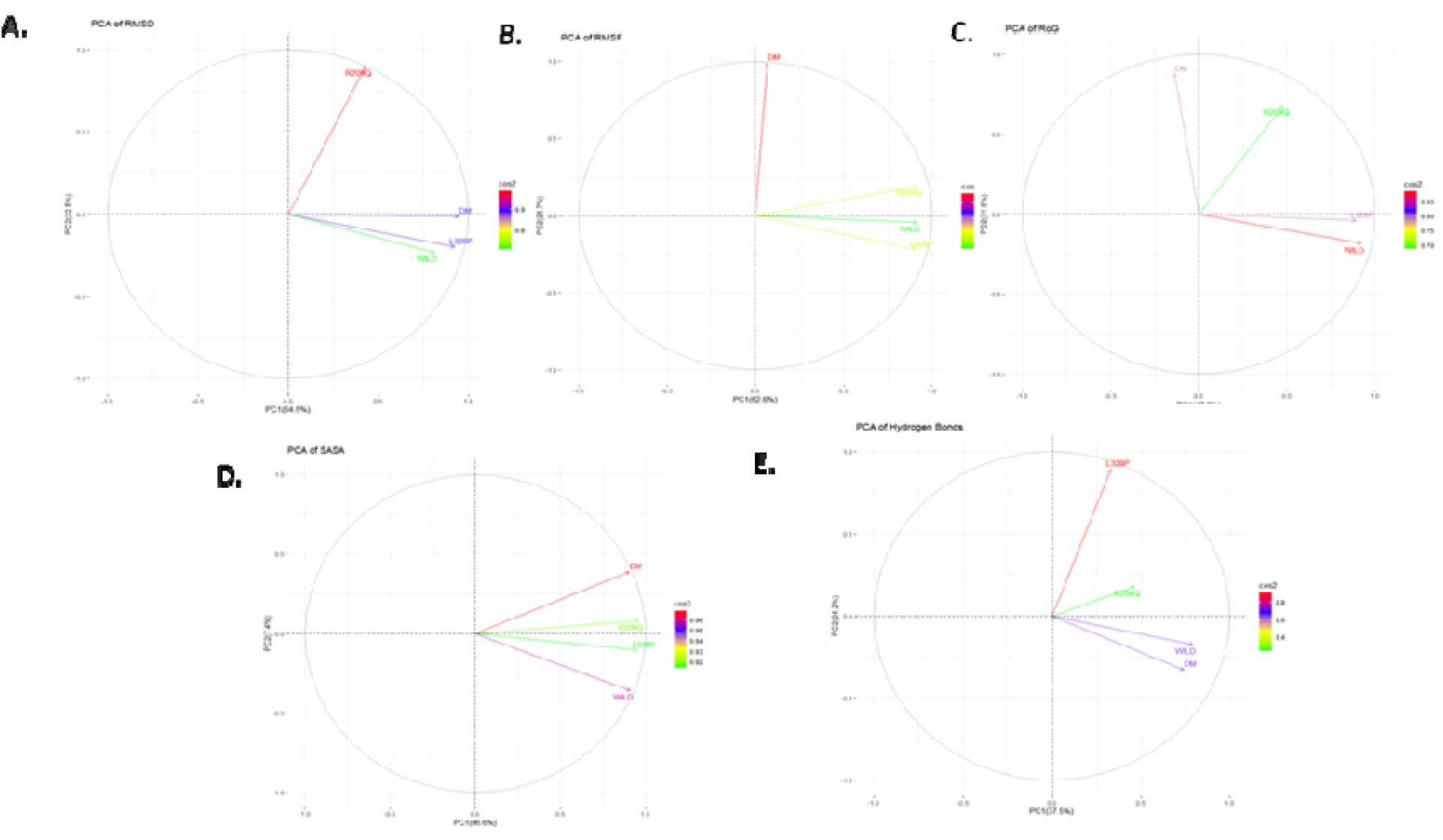
Principle Component Analysis (PCA) of GFAP variants Wild type- green, R209Q- yellow, L328P- blue and both L328P & R209Q- red A) RMSD B. RMSF C) Radius of gyration D.) Solvent Accessible Surface Area (SASA) E) Hydrogen bond

PTMs are essential to a variety of biological processes since they are present in practically all proteins and have an impact on their dynamics and structure. Numerous computational techniques have been developed for PTM analysis, demonstrating the usefulness of these instruments in anticipating changed areas that may be subjected to additional experimental investigation. We found functional alterations in P1, P2 and DM (Table 3).

In silico studies predicted that there is a significant change in the mutated protein structure compare to its wild type which might result in altered protein function and having effect on disease onset and progression. Hence, based on all the result the novel mutation (c. 983C>T) associated with another known mutation (c.626G>A) on GFAP gene transcribe to a mutant protein resulting to disease pathology. From the literature it was found that AxD is transmitted from one generation to the next in autosomal dominant pattern. However, in our study the mother is phenotypically normal even after having the missence variant (c. 626G>A), which was predicted to be disease causing by various prediction tools. These might indicating the concept of compound hetrozygosity, but further study is needed for understanding the disease pathophysiology.

## Conclusion

Type 1 Juvenile Alexander Disease is a type of Leukodystrophy, clinically characterized by seizure, spastic paresis, psychomotor retardation, milestone regression and pseudobulbular sign. Till to date mutation in GFAP gene is known to be the cause for AxD. In this study we screened a juvenile AxD with typical clinical feature and MRI finding with his parents. A novel (p.L328P) and a novel in Indian population (p.R209Q) GFAP variants in heterozygous condition were observed in the patient produces an altered GFAP protein product leading to formation of Rosenthal fibers, which impairs cell function and responsible for AxD disease phenotype.

## Data Availability

All data produced in the present study are available upon reasonable request to the authors
All data produced in the present work are contained in the manuscript

## Statement and Declarations

### Ethics declaration

This study was performed in line with the principles of the Declaration of Helsinki. Approval was granted by the Ethics Committee of Institute of Science, Banaras Hindu University (*Ref No: I*.*Sc/ECM-XII/2021-22/)*

### Conflict of Interest

The authors declare that there is no conflict of interest regarding publication of this article.

## Acknowledgement

The authors are grateful to the patient and his family members for their contribution to this study.

## Notes

### Competing Interest Statement

The authors have declared no competing interest.

### Funding Statement

This study did not receive any funding

### Author Declarations

Ethics Committee, Institute of Science, Banaras Hindu University, Varanasi, Ref No. I.Sc./ECM-XII/2021-22 Ethics committee, Institute of science, Banaras Hindu University, Varanasi, Uttar Pradesh, Ref No. I.Sc./ECM-XII/2021-22

